# Ratios Of Submitted Charge Amounts To Medicare Allowed Amounts Have Variably Worsened For Anesthesiologists And Certified Registered Nurse Anesthetists Serving Medicare Beneficiaries Across The United States During 2013-2021 Period

**DOI:** 10.1101/2024.01.17.24301422

**Authors:** Deepak Gupta, Yara Ismaeil

## Abstract

**Background:** Interstate variations among anesthesia providers in terms of their submitted charge amounts with corresponding Medicare allowed amounts have not been recently explored.

**Objective:** To quantify interstate variations in submitted charge amounts in relation to corresponding Medicare allowed amounts for the services provided by anesthesia providers during 2013-2021 for Medicare beneficiaries in the United States (U.S.).

**Materials and Methods:** Public Use Files for 2013-2021 period from Centers for Medicare & Medicaid Services were accessed to generate cumulative excel sheets of de-identified anesthesia providers data according to anesthesia providers stated location in fifty U.S. states and the District of Columbia. Thereafter, total number of providers, total number of services, total submitted charge amounts and total Medicare allowed amounts were finally tabulated according to the year of billed anesthesia service as well as according to anesthesia providers state.

**Results:** The submitted charge amount to Medicare allowed amount ratios varied from 4-9 across the states in 2013 for anesthesiologists as well as certified registered nurse anesthetists (CRNAs). These ratios worsened to 5-13 across the states for anesthesiologists in 2021 while these ratios worsened to 5-12 across the states for CRNAs in 2021. The greatest number of anesthesiologists serving Medicare beneficiaries have been located in California across the years while the greatest number of CRNAs serving Medicare beneficiaries have been located in Florida across the years. Overall, some states have more anesthesiologists per 100,000 population serving Medicare beneficiaries with District of Columbia having the maximum (28 anesthesiologists per 100,000 population) while some states have more CRNAs per 100,000 population serving Medicare beneficiaries with North Dakota having the maximum (38 CRNAs per 100,000 population).

**Conclusion:** Over the years (2013-2021), ratios of submitted charge amounts to Medicare allowed amounts have variably worsened for anesthesiologists and CRNAs serving Medicare beneficiaries across U.S. irrespective of the state stated by anesthesiologists and CRNAs as their primary location per National Plan and Provider Enumeration System.

## Introduction

The story of cost-to-charge ratios during billing by hospitals and other healthcare institutions is mesmerizing to the bewildered eyes while its understanding is enigmatic to the ignorant minds [1-2]. However, the vibrancy of system itself means that there may be more to this misunderstood but successful system than meets the bewildered eyes and ignorant minds. Interestingly, the cost may be zero or unquantified when it comes to billing by physicians and other providers. Therefore, charge-to-allowed ratios may be the only relevant ratios during billing by providers. However, charge-to-allowed ratios may be important during billing by healthcare institutions as well because their charge-to-allowed ratios may retroactively decide their cost-to-charge ratios as well.

After previously exploring a multi-year comparison of non-anesthesia providers with anesthesia providers namely anesthesiologists, certified registered nurse anesthetists (CRNAs) and anesthesiology assistants regarding variations in their submitted charge amounts with corresponding Medicare allowed amounts at the national level [3], it was deemed appropriate to explore interstate variations in submitted charge amounts with corresponding Medicare allowed amounts among anesthesia providers themselves.

Therefore, the primary purpose of this project was to quantify interstate variations in submitted charge amounts in relation to corresponding Medicare allowed amounts for the services provided by anesthesiologists, certified registered nurse anesthetists (CRNAs) and anesthesiology assistants during 2013-2021 for Medicare beneficiaries in the United States (U.S.).

## Materials and Methods

Institutional review board deemed this project as non-human participant research because of being based on Public Use Files of Centers for Medicare & Medicaid Services (CMS) [4-5]. Thereafter, CMS 2013-2021 Public Use Files were accessed after managing columns and applying advanced filters (See Raw Data Links as Supplementary PDF). Such data exports generated cumulative excel sheets of de-identified providers’ data according to their fifty U.S. states and the District of Columbia for anesthesiologists, CRNAs and anesthesiology assistants. Thereafter, the following data characteristics’ variations among anesthesiologists, CRNAs and anesthesiology assistants were tabulated for comparing according to their location (fifty U.S. states and the District of Columbia) as reported in National Plan and Provider Enumeration System (NPPES):

- Number of HCPCS
- Number of Medicare Beneficiaries
- Number of Services
- Total Submitted Charge Amount
- Total Medicare Allowed Amount
- Total Medicare Payment Amount
- Total Medicare Standardized Payment Amount
- Number of HCPCS Associated With Medical Services
- Number of Medicare Beneficiaries With Medical Services
- Number of Medical Services
- Total Medical Submitted Charge Amount
- Total Medical Medicare Allowed Amount
- Total Medical Medicare Payment Amount
- Total Medical Medicare Standardized Payment Amount

## Results

As anesthesiology assistants were not located in all fifty U.S. states and the District of Columbia, their data was not used for comparing with anesthesiologists and CRNAs. Moreover, for each year during the period 2013-2021, only number of providers, number of services, total submitted charge amounts and total Medicare allowed amounts were finally tabulated among anesthesiologists and CRNAs (See Raw Tables as Supplementary Excel Worksheet). Data points like number of HCPCS associated with medical services, number of Medicare beneficiaries with medical services, number of medical services, total medical submitted charge amount, total medical Medicare allowed amount, total medical Medicare payment amount, and total medical Medicare standardized payment amount were not present for all providers and thus excluded. Data points like number of HCPCS, number of Medicare beneficiaries, total Medicare payment amount, and total Medicare standardized payment amount were deemed irrelevant post-hoc for this project and thus excluded.

As detailed in Tables 1-3 derived from Raw Tables (Supplementary Excel Worksheet), the submitted charge amount to Medicare allowed amount ratios varied from 4-9 across the states in 2013 for anesthesiologists as well as CRNAs (Table 1). These ratios worsened to 5-13 across the states for anesthesiologists in 2021 while these ratios worsened to 5-12 across the states for CRNAs in 2021 (Table 3). As displayed in Map Video derived from Data for Maps (Supplementary Excel Worksheet), ratio of anesthesiologists to CRNAs, ratio of services per anesthesiologist to services per CRNA, and grand ratio of (ratio of submitted charge amount for anesthesiologist services to Medicare allowed amount for anesthesiologist services) to (ratio of submitted charge amount for CRNA services to Medicare allowed amount for CRNA services) varied across the states during 2013-2021 period. Moreover, anesthesiologists per 100,000 population and CRNAs per 100,000 population as derived for the year 2020 based on 2020 U.S. Census [6] varied across the states which is also displayed in Map Video as derived from Data for Maps (Supplementary Excel Worksheet).

**TABLE 1:** 2013-2015 Period Inter-State Variations In Submitted Charge Amounts By Medicare Allowed Amounts AND Services Billed Per Provider FOR Anesthesiologists & CRNAs Per Public Use Files From Centers for Medicare & Medicaid Services

| STATES | 2013 |  |  |  | 2014 |  |  |  | 2015 |  |  |  |
| --- | --- | --- | --- | --- | --- | --- | --- | --- | --- | --- | --- | --- |
|  | ANESTHESIOLOGISTS |  | CRNAS |  | ANESTHESIOLOGISTS |  | CRNAS |  | ANESTHESIOLOGISTS |  | CRNAS |  |
|  | CHARGE BY<br>ALLOWED RATIO | SERVICES PER<br>PROVIDER | CHARGE BY<br>ALLOWED RATIO | SERVICES PER<br>PROVIDER | CHARGE BY<br>ALLOWED RATIO | SERVICES PER<br>PROVIDER | CHARGE BY<br>ALLOWED RATIO | SERVICES PER<br>PROVIDER | CHARGE BY<br>ALLOWED RATIO | SERVICES PER<br>PROVIDER | CHARGE BY<br>ALLOWED RATIO | SERVICES PER<br>PROVIDER |
| Alaska | 5.01 | 202.10 | 4.07 | 100.57 | 4.76 | 484.44 | 3.93 | 94.02 | 4.90 | 618.70 | 4.07 | 100.09 |
| Alabama | 5.08 | 1073.02 | 5.55 | 204.46 | 5.14 | 1187.93 | 5.53 | 207.85 | 5.09 | 1215.05 | 5.78 | 214.81 |
| Arkansas | 4.00 | 632.37 | 3.98 | 243.52 | 4.15 | 731.62 | 4.21 | 241.13 | 4.40 | 843.40 | 4.53 | 241.65 |
| Arizona | 4.91 | 375.92 | 3.61 | 219.97 | 5.15 | 441.24 | 4.07 | 234.71 | 5.52 | 452.20 | 4.55 | 254.75 |
| California | 4.72 | 397.72 | 5.49 | 179.98 | 4.89 | 410.79 | 5.60 | 174.19 | 5.09 | 403.71 | 6.13 | 170.22 |
| Colorado | 6.64 | 262.06 | 6.20 | 154.19 | 6.61 | 270.31 | 6.32 | 157.52 | 6.85 | 298.20 | 6.65 | 164.99 |
| Connecticut | 7.16 | 377.12 | 6.08 | 192.04 | 7.59 | 411.83 | 6.98 | 194.03 | 7.53 | 375.01 | 7.71 | 196.31 |
| District of<br>Columbia | 5.45 | 265.40 | 4.83 | 155.31 | 5.72 | 268.94 | 5.98 | 164.42 | 5.98 | 305.17 | 6.22 | 170.27 |
| Delaware | 8.79 | 655.10 | 5.12 | 229.60 | 7.92 | 675.88 | 7.68 | 235.73 | 7.14 | 643.66 | 7.03 | 232.24 |
| Florida | 5.45 | 695.37 | 5.74 | 232.89 | 5.82 | 806.73 | 6.05 | 224.18 | 6.25 | 693.91 | 6.89 | 218.32 |
| Georgia | 5.81 | 803.27 | 5.34 | 200.06 | 5.82 | 1006.17 | 5.79 | 198.09 | 6.22 | 1043.75 | 6.81 | 211.01 |
| Hawaii | 5.11 | 206.86 | 6.05 | 141.45 | 5.11 | 252.16 | 7.02 | 152.09 | 5.00 | 242.11 | 6.35 | 213.06 |
| Iowa | 5.01 | 332.23 | 4.48 | 190.32 | 4.98 | 349.00 | 4.39 | 186.44 | 5.10 | 380.44 | 4.55 | 185.96 |
| Idaho | 4.40 | 298.53 | 4.62 | 127.56 | 4.43 | 399.54 | 4.65 | 134.62 | 5.37 | 548.43 | 5.04 | 137.96 |
| Illinois | 6.23 | 475.20 | 6.61 | 204.83 | 6.37 | 466.47 | 6.66 | 189.20 | 6.38 | 503.48 | 6.79 | 189.08 |
| Indiana | 5.17 | 433.50 | 5.61 | 205.99 | 5.19 | 443.07 | 5.66 | 219.42 | 5.23 | 583.41 | 5.91 | 225.24 |
| Kansas | 5.38 | 519.65 | 4.43 | 200.91 | 5.23 | 562.65 | 4.35 | 206.42 | 5.35 | 466.65 | 4.52 | 211.56 |
| Kentucky | 5.20 | 855.07 | 4.88 | 233.02 | 5.32 | 1057.20 | 4.91 | 224.88 | 5.35 | 1304.67 | 5.10 | 227.81 |
| Louisiana | 5.97 | 575.72 | 5.69 | 177.48 | 6.54 | 717.57 | 5.85 | 170.01 | 6.64 | 682.77 | 5.93 | 198.06 |
| Massachusetts | 6.38 | 420.33 | 6.08 | 201.63 | 6.35 | 427.51 | 6.08 | 212.08 | 6.44 | 443.71 | 6.35 | 216.90 |
| Maryland | 5.38 | 391.20 | 5.88 | 205.90 | 5.43 | 475.72 | 5.87 | 214.68 | 5.62 | 502.23 | 6.26 | 208.45 |
| Maine | 7.04 | 377.14 | 6.45 | 166.67 | 7.13 | 387.68 | 6.48 | 175.68 | 7.25 | 417.30 | 7.10 | 173.47 |
| Michigan | 6.73 | 660.62 | 6.14 | 188.80 | 7.13 | 663.22 | 6.30 | 179.87 | 7.66 | 616.84 | 6.75 | 180.13 |
| Minnesota | 6.26 | 273.71 | 5.43 | 88.29 | 6.49 | 280.27 | 5.34 | 85.42 | 6.68 | 294.93 | 5.54 | 86.07 |
| Missouri | 5.15 | 440.99 | 4.85 | 208.35 | 5.35 | 451.49 | 4.90 | 203.56 | 5.42 | 505.48 | 5.13 | 200.02 |
| Mississippi | 5.53 | 918.89 | 4.91 | 224.93 | 5.58 | 1074.79 | 5.05 | 231.34 | 5.55 | 1024.34 | 5.36 | 232.81 |
| Montana | 4.11 | 308.05 | 4.20 | 186.49 | 4.14 | 332.55 | 4.29 | 179.39 | 4.26 | 340.23 | 4.15 | 168.15 |
| North Carolina | 8.53 | 854.50 | 6.66 | 160.74 | 8.33 | 1013.88 | 6.87 | 148.32 | 8.97 | 765.98 | 7.28 | 166.82 |
| North Dakota | 5.96 | 378.58 | 4.60 | 136.81 | 6.48 | 374.06 | 4.73 | 137.28 | 6.50 | 412.08 | 5.22 | 131.27 |
| Nebraska | 4.34 | 478.38 | 4.36 | 221.01 | 4.71 | 374.28 | 4.25 | 217.09 | 4.73 | 380.88 | 4.51 | 214.75 |
| New Hampshire | 6.84 | 428.02 | 5.67 | 270.47 | 7.26 | 441.18 | 6.13 | 301.48 | 7.83 | 445.12 | 6.73 | 262.85 |
| New Jersey | 7.45 | 411.68 | 8.72 | 203.52 | 7.74 | 414.97 | 8.50 | 210.03 | 7.85 | 444.14 | 9.05 | 208.16 |
| New Mexico | 4.83 | 254.10 | 5.04 | 167.92 | 5.37 | 265.55 | 5.44 | 173.63 | 5.44 | 283.79 | 5.74 | 171.89 |
| Nevada | 5.30 | 434.39 | 5.01 | 213.51 | 5.49 | 474.57 | 4.75 | 201.70 | 5.95 | 480.70 | 5.11 | 211.39 |
| New York | 6.72 | 353.32 | 7.01 | 154.51 | 7.06 | 337.29 | 7.37 | 145.88 | 7.27 | 358.84 | 7.45 | 146.39 |
| Ohio | 5.23 | 505.41 | 4.95 | 150.40 | 5.46 | 506.19 | 4.98 | 149.39 | 5.63 | 510.43 | 5.44 | 148.03 |
| Oklahoma | 4.34 | 552.38 | 4.51 | 232.85 | 4.44 | 590.98 | 4.50 | 245.62 | 4.34 | 765.15 | 5.01 | 242.54 |
| Oregon | 5.08 | 215.63 | 4.93 | 150.27 | 5.18 | 217.62 | 4.82 | 149.18 | 5.33 | 216.99 | 5.25 | 147.24 |
| Pennsylvania | 6.72 | 428.58 | 6.51 | 146.68 | 6.75 | 451.42 | 6.44 | 148.23 | 6.91 | 502.53 | 6.83 | 149.91 |
| Rhode Island | 5.69 | 309.73 | 5.07 | 162.20 | 6.03 | 317.23 | 4.96 | 163.54 | 6.38 | 330.31 | 5.53 | 159.66 |
| South Carolina | 7.17 | 917.93 | 7.34 | 191.18 | 7.55 | 905.40 | 7.21 | 187.16 | 7.67 | 891.55 | 7.69 | 185.51 |
| South Dakota | 5.13 | 687.19 | 7.08 | 144.86 | 4.90 | 676.44 | 7.22 | 150.14 | 5.46 | 641.58 | 7.56 | 145.16 |
| Tennessee | 6.19 | 733.04 | 5.80 | 192.81 | 6.37 | 760.84 | 6.02 | 191.86 | 6.88 | 656.65 | 6.53 | 203.96 |
| Texas | 5.99 | 548.40 | 6.56 | 194.75 | 6.60 | 567.67 | 7.07 | 191.89 | 7.19 | 567.62 | 7.84 | 195.66 |
| Utah | 4.29 | 356.07 | 3.85 | 169.74 | 4.33 | 328.19 | 3.93 | 163.44 | 4.39 | 350.59 | 4.36 | 165.69 |
| Virginia | 7.29 | 446.11 | 5.87 | 192.62 | 7.53 | 448.64 | 6.12 | 192.43 | 7.92 | 457.82 | 7.25 | 216.95 |
| Vermont | 6.54 | 402.44 | 6.00 | 134.50 | 6.12 | 624.72 | 6.04 | 148.03 | 5.95 | 567.00 | 5.73 | 162.53 |
| Washington | 4.77 | 237.78 | 4.71 | 176.96 | 4.80 | 241.12 | 4.86 | 176.56 | 5.06 | 241.82 | 5.03 | 178.37 |
| Wisconsin | 7.08 | 279.12 | 8.35 | 124.61 | 8.75 | 288.94 | 8.73 | 127.22 | 9.56 | 286.47 | 9.22 | 133.52 |
| West Virginia | 5.59 | 712.12 | 4.75 | 176.50 | 5.92 | 527.04 | 4.68 | 167.57 | 6.02 | 506.00 | 4.87 | 173.32 |
| Wyoming | 3.95 | 270.69 | 4.19 | 150.76 | 4.61 | 251.87 | 4.34 | 126.05 | 4.67 | 253.13 | 4.43 | 133.59 |
| UNITED STATES | 5.92 | 483.36 | 5.79 | 184.22 | 6.15 | 515.33 | 5.97 | 181.93 | 6.40 | 519.27 | 6.44 | 185.95 |

Sourced from Data provided by Centers for Medicare & Medicaid Services Public Use Files

**TABLE 2:** 2016-2018 Period Inter-State Variations In Submitted Charge Amounts By Medicare Allowed Amounts AND Services Billed Per Provider FOR Anesthesiologists & CRNAs Per Public Use Files From Centers for Medicare & Medicaid Services

| STATES | 2016 |  |  |  | 2017 |  |  |  | 2018 |  |  |  |
| --- | --- | --- | --- | --- | --- | --- | --- | --- | --- | --- | --- | --- |
|  | ANESTHESIOLOGISTS |  | CRNAS |  | ANESTHESIOLOGISTS |  | CRNAS |  | ANESTHESIOLOGISTS |  | CRNAS |  |
|  | CHARGE BY<br>ALLOWED RATIO | SERVICES PER<br>PROVIDER | CHARGE BY<br>ALLOWED RATIO | SERVICES PER<br>PROVIDER | CHARGE BY<br>ALLOWED RATIO | SERVICES PER<br>PROVIDER | CHARGE BY<br>ALLOWED RATIO | SERVICES PER<br>PROVIDER | CHARGE BY<br>ALLOWED RATIO | SERVICES PER<br>PROVIDER | CHARGE BY<br>ALLOWED RATIO | SERVICES PER<br>PROVIDER |
| Alaska | 5.68 | 362.01 | 4.35 | 109.85 | 6.09 | 425.17 | 4.88 | 119.14 | 6.45 | 569.88 | 5.17 | 118.22 |
| Alabama | 5.33 | 1224.63 | 6.27 | 216.17 | 5.87 | 1014.78 | 6.64 | 186.55 | 5.97 | 910.63 | 6.97 | 174.75 |
| Arkansas | 4.77 | 707.01 | 4.87 | 242.00 | 4.88 | 692.88 | 5.11 | 236.32 | 4.67 | 742.64 | 5.13 | 240.49 |
| Arizona | 6.46 | 411.05 | 5.39 | 275.06 | 7.33 | 417.06 | 6.13 | 325.02 | 7.93 | 452.09 | 8.07 | 315.22 |
| California | 5.42 | 415.49 | 6.77 | 175.55 | 5.66 | 416.87 | 7.08 | 179.19 | 5.94 | 421.04 | 7.38 | 180.19 |
| Colorado | 7.16 | 383.23 | 7.10 | 170.60 | 7.69 | 412.10 | 7.29 | 164.62 | 8.10 | 388.93 | 7.42 | 173.09 |
| Connecticut | 7.88 | 369.83 | 8.21 | 194.38 | 8.73 | 376.79 | 8.95 | 186.61 | 9.26 | 348.56 | 9.74 | 166.97 |
| District of<br>Columbia | 6.19 | 301.62 | 5.77 | 179.16 | 6.21 | 334.33 | 6.55 | 195.60 | 7.55 | 338.36 | 7.49 | 179.89 |
| Delaware | 7.91 | 655.60 | 7.76 | 234.34 | 8.00 | 633.17 | 8.19 | 231.01 | 7.90 | 761.67 | 8.63 | 225.50 |
| Florida | 6.90 | 653.26 | 8.01 | 218.09 | 7.23 | 691.90 | 8.35 | 208.68 | 7.60 | 651.38 | 8.77 | 205.60 |
| Georgia | 7.23 | 777.66 | 7.88 | 236.75 | 7.48 | 736.63 | 8.43 | 242.48 | 7.60 | 737.92 | 8.61 | 195.81 |
| Hawaii | 5.26 | 252.07 | 6.53 | 224.24 | 5.35 | 288.07 | 7.18 | 203.89 | 5.48 | 275.50 | 7.86 | 220.39 |
| Iowa | 5.53 | 386.60 | 4.73 | 185.57 | 5.83 | 373.67 | 4.89 | 181.26 | 6.34 | 375.73 | 5.39 | 187.12 |
| Idaho | 5.48 | 331.05 | 5.30 | 157.32 | 4.96 | 314.29 | 5.38 | 157.90 | 5.25 | 323.09 | 5.36 | 161.31 |
| Illinois | 7.03 | 508.01 | 7.58 | 208.73 | 7.44 | 479.23 | 7.95 | 215.39 | 8.01 | 509.88 | 8.54 | 224.17 |
| Indiana | 5.60 | 421.10 | 6.35 | 232.99 | 5.63 | 419.67 | 6.68 | 255.07 | 5.94 | 471.00 | 7.19 | 243.35 |
| Kansas | 5.52 | 484.12 | 4.85 | 213.62 | 5.78 | 464.21 | 5.08 | 210.80 | 5.79 | 458.92 | 5.24 | 210.17 |
| Kentucky | 5.60 | 1073.02 | 5.55 | 239.23 | 5.71 | 1123.65 | 5.74 | 239.33 | 5.90 | 1186.49 | 6.19 | 241.02 |
| Louisiana | 7.07 | 554.78 | 6.17 | 210.29 | 7.19 | 544.78 | 6.30 | 208.85 | 7.06 | 557.57 | 6.36 | 201.45 |
| Massachusetts | 6.65 | 481.10 | 6.86 | 227.07 | 6.69 | 482.27 | 6.97 | 217.27 | 6.66 | 516.77 | 7.00 | 213.94 |
| Maryland | 6.42 | 416.08 | 6.86 | 211.75 | 6.58 | 475.92 | 6.98 | 208.01 | 6.80 | 505.57 | 7.65 | 207.92 |
| Maine | 7.65 | 409.16 | 7.48 | 165.99 | 7.55 | 409.92 | 7.57 | 169.67 | 7.81 | 372.18 | 8.12 | 157.48 |
| Michigan | 8.35 | 607.78 | 7.61 | 178.15 | 8.87 | 608.34 | 7.82 | 172.58 | 9.06 | 637.54 | 8.51 | 163.58 |
| Minnesota | 6.89 | 300.94 | 5.92 | 88.07 | 7.28 | 296.45 | 5.98 | 87.21 | 8.00 | 274.67 | 6.53 | 88.08 |
| Missouri | 5.66 | 530.33 | 5.62 | 208.79 | 5.90 | 496.45 | 5.71 | 206.56 | 6.46 | 476.80 | 5.90 | 194.78 |
| Mississippi | 6.08 | 689.59 | 5.58 | 235.38 | 6.47 | 696.81 | 5.75 | 254.42 | 6.94 | 601.89 | 5.82 | 264.47 |
| Montana | 4.36 | 312.80 | 4.53 | 177.64 | 4.43 | 326.39 | 4.78 | 177.76 | 4.31 | 324.54 | 4.61 | 202.06 |
| North Carolina | 9.46 | 688.21 | 7.46 | 158.66 | 9.74 | 716.72 | 7.69 | 159.76 | 9.84 | 696.05 | 7.90 | 157.14 |
| North Dakota | 6.73 | 405.56 | 5.63 | 127.65 | 6.92 | 446.44 | 6.03 | 133.32 | 7.01 | 463.77 | 6.32 | 136.51 |
| Nebraska | 4.81 | 443.00 | 4.70 | 217.54 | 4.95 | 394.26 | 4.96 | 228.49 | 5.00 | 371.98 | 5.01 | 215.51 |
| New Hampshire | 8.59 | 457.07 | 8.22 | 268.38 | 8.76 | 481.33 | 9.21 | 241.62 | 9.40 | 456.87 | 9.99 | 224.36 |
| New Jersey | 8.31 | 459.70 | 8.96 | 199.15 | 8.88 | 453.73 | 9.18 | 193.84 | 9.22 | 447.10 | 9.87 | 189.56 |
| New Mexico | 5.66 | 306.63 | 5.82 | 177.01 | 5.88 | 335.10 | 6.08 | 183.52 | 5.78 | 383.34 | 6.51 | 176.37 |
| Nevada | 6.15 | 438.64 | 5.66 | 198.48 | 6.90 | 480.93 | 5.86 | 222.22 | 7.65 | 491.62 | 6.45 | 219.11 |
| New York | 7.80 | 372.23 | 7.65 | 151.45 | 8.25 | 384.38 | 8.02 | 151.91 | 9.00 | 391.74 | 8.77 | 153.95 |
| Ohio | 5.97 | 482.25 | 5.98 | 162.26 | 6.06 | 465.27 | 6.10 | 159.51 | 6.42 | 429.45 | 6.67 | 157.36 |
| Oklahoma | 4.43 | 720.51 | 5.57 | 258.05 | 4.50 | 700.72 | 5.56 | 266.20 | 5.08 | 732.84 | 6.19 | 262.17 |
| Oregon | 5.12 | 216.14 | 5.45 | 159.00 | 5.64 | 223.66 | 5.79 | 165.11 | 5.88 | 231.23 | 5.79 | 166.55 |
| Pennsylvania | 7.28 | 499.50 | 7.38 | 156.96 | 7.48 | 499.98 | 7.47 | 152.38 | 7.72 | 513.93 | 7.85 | 153.74 |
| Rhode Island | 6.67 | 333.21 | 6.05 | 168.04 | 7.03 | 294.85 | 7.16 | 154.73 | 7.01 | 338.49 | 7.36 | 156.09 |
| South Carolina | 8.75 | 852.75 | 8.18 | 189.60 | 8.73 | 1137.73 | 8.41 | 183.91 | 8.45 | 1328.70 | 8.68 | 181.46 |
| South Dakota | 5.55 | 659.19 | 7.62 | 153.04 | 5.37 | 754.12 | 7.17 | 165.93 | 5.23 | 790.56 | 7.97 | 172.16 |
| Tennessee | 6.64 | 709.34 | 7.04 | 201.21 | 6.13 | 959.96 | 7.17 | 198.00 | 7.21 | 763.87 | 7.42 | 185.98 |
| Texas | 8.07 | 476.39 | 8.49 | 195.74 | 8.43 | 486.58 | 8.93 | 200.62 | 8.95 | 503.43 | 9.61 | 193.75 |
| Utah | 4.61 | 369.31 | 4.90 | 166.37 | 4.63 | 393.46 | 5.01 | 169.84 | 4.60 | 418.33 | 5.03 | 169.09 |
| Virginia | 8.54 | 504.56 | 7.57 | 204.18 | 8.78 | 538.90 | 7.69 | 205.33 | 8.86 | 597.30 | 8.25 | 216.47 |
| Vermont | 6.20 | 603.04 | 5.88 | 180.00 | 5.88 | 630.38 | 5.82 | 172.41 | 5.81 | 492.56 | 6.19 | 170.20 |
| Washington | 5.36 | 252.59 | 5.22 | 186.51 | 5.53 | 270.54 | 5.36 | 186.28 | 5.72 | 289.99 | 5.56 | 190.88 |
| Wisconsin | 10.30 | 284.86 | 9.70 | 136.02 | 10.72 | 283.37 | 10.14 | 136.80 | 11.28 | 287.85 | 10.79 | 140.69 |
| West Virginia | 6.51 | 526.20 | 5.47 | 166.48 | 7.86 | 451.25 | 6.63 | 163.27 | 8.69 | 456.33 | 7.68 | 156.84 |
| Wyoming | 4.53 | 257.61 | 4.73 | 149.64 | 5.54 | 313.35 | 5.08 | 147.30 | 5.89 | 362.20 | 5.52 | 148.13 |
| **UNITED STATES** | **6.88** | **492.79** | **6.99** | **189.82** | **7.18** | **503.72** | **7.26** | **188.45** | **7.53** | **508.26** | **7.69** | **184.69** |

Sourced from Data provided by Centers for Medicare & Medicaid Services Public Use Files

**TABLE 3:** 2019-2021 Period Inter-State Variations In Submitted Charge Amounts By Medicare Allowed Amounts AND Services Billed Per Provider FOR Anesthesiologists & CRNAs Per Public Use Files From Centers for Medicare & Medicaid Services

| STATES | 2019 |  |  |  | 2020 |  |  |  | 2021 |  |  |  |
| --- | --- | --- | --- | --- | --- | --- | --- | --- | --- | --- | --- | --- |
|  | ANESTHESIOLOGISTS |  | CRNAS |  | ANESTHESIOLOGISTS |  | CRNAS |  | ANESTHESIOLOGISTS |  | CRNAS |  |
|  | CHARGE BY<br>ALLOWED RATIO | SERVICES PER<br>PROVIDER | CHARGE BY<br>ALLOWED RATIO | SERVICES PER<br>PROVIDER | CHARGE BY<br>ALLOWED RATIO | SERVICES PER<br>PROVIDER | CHARGE BY<br>ALLOWED RATIO | SERVICES PER<br>PROVIDER | CHARGE BY<br>ALLOWED RATIO | SERVICES PER<br>PROVIDER | CHARGE BY<br>ALLOWED RATIO | SERVICES PER<br>PROVIDER |
| Alaska | 6.81 | 702.75 | 5.28 | 130.41 | 6.44 | 663.69 | 5.39 | 114.29 | 6.69 | 732.63 | 5.50 | 136.62 |
| Alabama | 6.20 | 843.19 | 7.08 | 168.32 | 5.92 | 726.56 | 7.06 | 137.68 | 6.20 | 711.54 | 7.53 | 137.92 |
| Arkansas | 5.10 | 647.11 | 5.03 | 240.92 | 5.04 | 613.27 | 5.21 | 206.60 | 5.23 | 642.36 | 5.33 | 221.54 |
| Arizona | 8.39 | 475.67 | 9.85 | 326.25 | 8.31 | 440.90 | 9.31 | 260.67 | 8.07 | 495.03 | 9.92 | 254.49 |
| California | 6.09 | 414.30 | 7.82 | 176.55 | 6.16 | 340.92 | 8.43 | 153.72 | 6.36 | 379.24 | 9.11 | 171.94 |
| Colorado | 8.43 | 328.24 | 7.56 | 165.46 | 8.54 | 291.42 | 7.60 | 138.57 | 8.72 | 334.10 | 8.13 | 151.39 |
| Connecticut | 9.19 | 379.67 | 9.85 | 155.84 | 9.00 | 333.39 | 9.93 | 117.56 | 9.60 | 406.60 | 10.83 | 129.24 |
| District of<br>Columbia | 7.36 | 356.77 | 8.58 | 169.69 | 7.54 | 343.05 | 8.64 | 138.03 | 7.31 | 365.13 | 8.52 | 177.61 |
| Delaware | 7.77 | 757.08 | 8.39 | 227.60 | 7.33 | 663.56 | 8.47 | 189.01 | 8.23 | 650.18 | 9.16 | 214.93 |
| Florida | 7.90 | 622.73 | 9.01 | 201.48 | 7.85 | 526.54 | 9.20 | 164.21 | 7.72 | 513.97 | 9.65 | 171.21 |
| Georgia | 7.72 | 690.60 | 8.72 | 194.75 | 7.75 | 556.01 | 9.14 | 159.72 | 7.88 | 553.87 | 9.81 | 159.92 |
| Hawaii | 5.91 | 267.21 | 8.17 | 206.56 | 5.99 | 232.88 | 9.63 | 143.44 | 6.22 | 242.83 | 11.97 | 177.69 |
| Iowa | 6.38 | 370.40 | 5.52 | 192.69 | 6.48 | 302.65 | 5.59 | 163.64 | 6.63 | 344.08 | 6.00 | 187.50 |
| Idaho | 5.29 | 335.11 | 5.55 | 165.81 | 5.35 | 337.40 | 5.67 | 147.26 | 5.50 | 404.24 | 5.76 | 158.72 |
| Illinois | 8.46 | 519.87 | 9.01 | 248.97 | 8.58 | 480.08 | 9.50 | 213.78 | 8.85 | 549.61 | 10.22 | 230.16 |
| Indiana | 6.08 | 449.05 | 7.20 | 245.82 | 6.31 | 363.95 | 7.22 | 202.86 | 6.68 | 388.89 | 8.06 | 204.16 |
| Kansas | 5.83 | 538.73 | 5.23 | 235.32 | 5.65 | 420.94 | 5.33 | 204.82 | 5.90 | 446.30 | 5.66 | 215.19 |
| Kentucky | 6.05 | 1061.15 | 6.30 | 225.34 | 5.84 | 875.40 | 6.61 | 176.20 | 6.21 | 750.23 | 7.27 | 172.12 |
| Louisiana | 7.20 | 503.50 | 6.43 | 181.72 | 7.04 | 437.67 | 6.55 | 141.29 | 7.22 | 443.75 | 7.00 | 145.11 |
| Massachusetts | 6.73 | 536.88 | 7.47 | 216.93 | 6.77 | 440.30 | 7.62 | 167.50 | 6.98 | 462.75 | 7.77 | 196.52 |
| Maryland | 7.28 | 457.42 | 8.21 | 221.55 | 7.06 | 403.54 | 8.49 | 177.47 | 7.18 | 426.32 | 8.97 | 195.47 |
| Maine | 7.89 | 364.11 | 8.71 | 158.07 | 8.01 | 287.14 | 8.72 | 121.18 | 8.01 | 287.93 | 9.40 | 122.98 |
| Michigan | 9.56 | 627.00 | 9.09 | 163.00 | 9.87 | 472.80 | 9.39 | 122.49 | 10.70 | 487.52 | 11.36 | 129.11 |
| Minnesota | 7.45 | 321.44 | 6.34 | 106.25 | 6.97 | 279.84 | 6.48 | 91.20 | 7.11 | 293.43 | 6.45 | 94.11 |
| Missouri | 6.72 | 435.53 | 6.02 | 189.75 | 6.83 | 353.86 | 6.38 | 156.21 | 7.15 | 359.73 | 6.95 | 170.06 |
| Mississippi | 7.21 | 621.78 | 6.00 | 262.88 | 7.00 | 563.80 | 5.92 | 214.68 | 6.85 | 577.31 | 6.00 | 227.89 |
| Montana | 4.38 | 315.42 | 4.94 | 202.82 | 4.73 | 292.99 | 4.83 | 172.00 | 5.13 | 302.78 | 6.07 | 206.62 |
| North Carolina | 9.88 | 704.76 | 7.93 | 158.40 | 9.89 | 609.53 | 8.23 | 132.26 | 10.10 | 597.75 | 8.58 | 134.62 |
| North Dakota | 8.23 | 423.06 | 6.58 | 148.45 | 7.92 | 367.11 | 6.61 | 137.62 | 8.51 | 410.10 | 6.89 | 163.02 |
| Nebraska | 5.09 | 327.84 | 5.17 | 218.30 | 5.23 | 318.09 | 5.30 | 191.44 | 5.71 | 355.95 | 6.30 | 202.12 |
| New Hampshire | 10.29 | 446.81 | 10.48 | 211.00 | 10.15 | 355.08 | 10.80 | 152.92 | 10.15 | 425.00 | 11.54 | 162.29 |
| New Jersey | 9.31 | 422.84 | 10.03 | 175.13 | 9.23 | 320.23 | 10.33 | 137.00 | 9.87 | 373.48 | 11.19 | 152.31 |
| New Mexico | 6.25 | 403.52 | 6.80 | 197.92 | 6.24 | 338.46 | 7.05 | 150.67 | 6.60 | 445.58 | 7.62 | 177.63 |
| Nevada | 7.75 | 507.01 | 7.27 | 214.46 | 7.85 | 469.59 | 7.58 | 202.64 | 8.68 | 441.07 | 7.92 | 214.85 |
| New York | 9.51 | 454.28 | 9.10 | 150.08 | 9.69 | 371.82 | 9.99 | 120.05 | 10.50 | 431.52 | 11.42 | 133.10 |
| Ohio | 6.53 | 413.20 | 6.92 | 156.94 | 6.57 | 343.88 | 7.03 | 131.89 | 6.88 | 350.96 | 7.27 | 140.32 |
| Oklahoma | 5.07 | 925.71 | 6.19 | 266.77 | 5.07 | 754.32 | 6.54 | 225.06 | 4.94 | 819.60 | 6.85 | 251.77 |
| Oregon | 6.02 | 229.39 | 6.29 | 209.13 | 5.81 | 198.89 | 6.47 | 188.03 | 5.91 | 209.02 | 6.26 | 173.19 |
| Pennsylvania | 7.74 | 477.34 | 8.05 | 154.55 | 7.84 | 401.67 | 8.24 | 125.31 | 7.98 | 481.59 | 8.62 | 135.55 |
| Rhode Island | 7.44 | 364.94 | 8.03 | 164.88 | 7.57 | 317.77 | 8.00 | 135.58 | 7.36 | 387.37 | 8.20 | 172.18 |
| South Carolina | 9.12 | 967.84 | 8.84 | 188.19 | 9.00 | 831.94 | 9.03 | 155.80 | 8.68 | 990.44 | 9.13 | 156.51 |
| South Dakota | 5.21 | 804.84 | 8.28 | 170.94 | 5.58 | 645.30 | 8.49 | 150.40 | 5.67 | 740.47 | 9.31 | 164.03 |
| Tennessee | 7.58 | 572.28 | 7.48 | 180.28 | 7.65 | 465.26 | 7.60 | 149.90 | 7.74 | 465.58 | 7.98 | 156.16 |
| Texas | 9.24 | 490.99 | 10.04 | 191.26 | 9.42 | 443.26 | 10.27 | 162.68 | 9.44 | 475.23 | 11.02 | 179.38 |
| Utah | 4.58 | 444.73 | 5.29 | 171.81 | 4.79 | 389.26 | 5.64 | 161.05 | 5.23 | 401.89 | 6.20 | 208.58 |
| Virginia | 8.97 | 678.93 | 8.93 | 213.34 | 9.43 | 563.47 | 8.94 | 174.77 | 9.61 | 631.07 | 8.72 | 181.53 |
| Vermont | 6.16 | 432.86 | 6.03 | 168.68 | 6.34 | 262.66 | 6.25 | 139.18 | 6.28 | 291.25 | 6.71 | 164.71 |
| Washington | 5.96 | 299.06 | 5.73 | 198.84 | 5.99 | 270.18 | 6.19 | 166.32 | 6.26 | 291.17 | 6.68 | 168.86 |
| Wisconsin | 11.87 | 272.99 | 11.24 | 138.34 | 12.31 | 229.65 | 11.77 | 120.11 | 13.07 | 244.67 | 12.48 | 130.42 |
| West Virginia | 9.73 | 461.14 | 8.52 | 154.65 | 9.82 | 392.41 | 8.72 | 125.74 | 10.11 | 395.99 | 9.68 | 130.40 |
| Wyoming | 5.96 | 324.64 | 6.25 | 163.89 | 6.17 | 334.03 | 6.35 | 146.32 | 6.90 | 337.77 | 6.64 | 177.26 |
| UNITED STATES | 7.77 | 497.65 | 7.95 | 184.92 | 7.81 | 420.79 | 8.16 | 152.47 | 8.07 | 452.23 | 8.70 | 162.25 |

Sourced from Data provided by Centers for Medicare & Medicaid Services Public Use Files

## Discussion

There were few interesting findings for anesthesiologists and CRNAs located in various states. First and foremost, the submitted charge amount to Medicare allowed amount ratios are worsening over the years indicating that submitted charge amounts by them may have increased and/or Medicare allowed amounts may have decreased over the years while the grand ratios varying from 1-2 implicating that both anesthesiologists and CRNAs had similar worsening in the submitted charge amount to Medicare allowed amount ratios according to the states they were located in. Secondly, services per provider variations across the states may be reflecting the preponderance of served Medicare beneficiaries in some states over others while medical directioning and medical supervision models may be the underlying reason for number of services per anesthesiologist overshooting number of services per CRNA in the ratios of 1-7 in every state. Thirdly, the greatest number of anesthesiologists serving Medicare beneficiaries have been located in California across the years while the greatest number of CRNAs serving Medicare beneficiaries have been located in Florida across the years thus most likely reflecting variations in state taxation policies, variations in state living costs and variations in ease of interstate travel according to various states used as their primary locations. Last but not least, anesthesiologists to CRNAs ratio varied from 0-7 across the states over the years meaning that some states have more anesthesiologists per 100,000 population serving Medicare beneficiaries with District of Columbia having the maximum (28 anesthesiologists per 100,000 population) while some states have more CRNAs per 100,000 population serving Medicare beneficiaries with North Dakota having the maximum (38 CRNAs per 100,000 population).

There were few limitations to this project. It could not be logistically demarcated within CMS data whether anesthesiologists and CRNAs were billing only for anesthesia services or also for pain services, critical care services, or any other non-anesthesia services under their purview. As the location of provider (anesthesiologist or CRNA) may not be the same as location of Medicare beneficiary, these interstate variations based on providers’ location may not be reflecting the interstate variations based on served Medicare beneficiaries’ location. Moreover, even providers’ location may not have been updated in NPPES annually [7] considering the mobility of providers especially anesthesiologists and CRNAs and their potential to serve more than one state’s Medicare beneficiaries across the years. Therefore, this project’s interstate variations’ tabulations may be giving only a skewed insight while the actual interstate variations into submitted charge amounts and Medicare allowed amounts can only be determined when researchers may apply for access to CMS data as delineated according to Medicare beneficiaries’ state in non-Public Use Files (Research Identifiable Files and Limited Data Sets [8]) rather than according to providers’ state in Public Use Files.

## Conclusion

Over the years (2013-2021), ratios of submitted charge amounts to Medicare allowed amounts have variably worsened for anesthesiologists and CRNAs serving Medicare beneficiaries across U.S. irrespective of the state stated by anesthesiologists and CRNAs as their primary location per NPPES.

## Supporting information

RAW TABLES

DATA FOR MAPS

RAW DATA LINKS

MAP VIDEO

IRB non-HPR

## Data Availability

All data produced in the present work are contained in the manuscript with links how to access raw data at public use files from CMS contained in the supplementary files

## Acknowledgement

The authors are indebted to Centers for Medicare & Medicaid Services for their 2013-2021 Public Use Files and 2020 U.S. Census for their public domain data (Courtesy U.S. Government) so that this project can come to fruition.

