## Supplementary material for "Ratios Of Submitted Charge Amounts To Medicare Allowed Amounts Have Variably Worsened For Anesthesiologists And Certified Registered Nurse Anesthetists Serving Medicare Beneficiaries Across The United States During 2013-2021 Period": RAW DATA LINKS

2013

https://data.cms.gov/provider-summary-by-type-of-service/medicare-physician-other-practitioners/medicare-physician-other-practitioners-by-provider/data/2013?query=%7B%22filters%22%3A%7B%22list%22%3A%5B%7B%22conjunction%22%3A%7B%22value%22%3A%22OR%22%7D%2C%22conditions%22%3A%5B%7B%22column%22%3A%7B%22value%22%3A%22Rndrng_Prvdr_Type%22%7D%2C%22comparator%22%3A%7B%22value%22%3A%22%3D%22%7D%2C%22filterValue%22%3A%5B%22anesthesiology%22%5D%7D%2C%7B%22column%22%3A%7B%22value%22%3A%22Rndrng_Prvdr_Type%22%7D%2C%22comparator%22%3A%7B%22value%22%3A%22%3D%22%7D%2C%22filterValue%22%3A%5B%22CRNA%22%5D%7D%2C%7B%22column%22%3A%7B%22value%22%3A%22Rndrng_Prvdr_Type%22%7D%2C%22comparator%22%3A%7B%22value%22%3A%22%3D%22%7D%2C%22filterValue%22%3A%5B%22anesthesiologist%20assistants%22%5D%7D%5D%7D%5D%2C%22rootConjunction%22%3A%7B%22value%22%3A%22AND%22%7D%7D%2C%22keywords%22%3A%22%22%2C%22offset%22%3A0%2C%22limit%22%3A10%2C%22sort%22%3A%7B%22sortBy%22%3Anull%2C%22sortOrder%22%3Anull%7D%2C%22columns%22%3A%5B%22Rndrng_Prvdr_Type%22%2C%22Rndrng_Prvdr_State_Abrvtn%22%2C%22Tot_HCPCS_Cds%22%2C%22Tot_Benes%22%2C%22Tot_Srvcs%22%2C%22Tot_Sbmtd_Chrg%22%2C%22Tot_Mdcr_Alowd_Amt%22%2C%22Tot_Mdcr_Pymt_Amt%22%2C%22Tot_Mdcr_Stdzd_Amt%22%2C%22Med_Tot_HCPCS_Cds%22%2C%22Med_Tot_Benes%22%2C%22Med_Tot_Srvcs%22%2C%22Med_Sbmtd_Chrg%22%2C%22Med_Mdcr_Alowd_Amt%22%2C%22Med_Mdcr_Pymt_Amt%22%2C%22Med_Mdcr_Stdzd_Amt%22%5D%7D

2014

https://data.cms.gov/provider-summary-by-type-of-service/medicare-physician-other-practitioners/medicare-physician-other-practitioners-by-provider/data/2014?query=%7B%22filters%22%3A%7B%22list%22%3A%5B%7B%22conjunction%22%3A%7B%22value%22%3A%22OR%22%7D%2C%22conditions%22%3A%5B%7B%22column%22%3A%7B%22value%22%3A%22Rndrng_Prvdr_Type%22%7D%2C%22comparator%22%3A%7B%22value%22%3A%22%3D%22%7D%2C%22filterValue%22%3A%5B%22anesthesiology%22%5D%7D%2C%7B%22column%22%3A%7B%22value%22%3A%22Rndrng_Prvdr_Type%22%7D%2C%22comparator%22%3A%7B%22value%22%3A%22%3D%22%7D%2C%22filterValue%22%3A%5B%22CRNA%22%5D%7D%2C%7B%22column%22%3A%7B%22value%22%3A%22Rndrng_Prvdr_Type%22%7D%2C%22comparator%22%3A%7B%22value%22%3A%22%3D%22%7D%2C%22filterValue%22%3A%5B%22anesthesiologist%20assistants%22%5D%7D%5D%7D%5D%2C%22rootConjunction%22%3A%7B%22value%22%3A%22AND%22%7D%7D%2C%22keywords%22%3A%22%22%2C%22offset%22%3A0%2C%22limit%22%3A10%2C%22sort%22%3A%7B%22sortBy%22%3Anull%2C%22sortOrder%22%3Anull%7D%2C%22columns%22%3A%5B%22Rndrng_Prvdr_Type%22%2C%22Rndrng_Prvdr_State_Abrvtn%22%2C%22Tot_HCPCS_Cds%22%2C%22Tot_Benes%22%2C%22Tot_Srvcs%22%2C%22Tot_Sbmtd_Chrg%22%2C%22Tot_Mdcr_Alowd_Amt%22%2C%22Tot_Mdcr_Pymt_Amt%22%2C%22Tot_Mdcr_Stdzd_Amt%22%2C%22Med_Tot_HCPCS_Cds%22%2C%22Med_Tot_Benes%22%2C%22Med_Tot_Srvcs%22%2C%22Med_Sbmtd_Chrg%22%2C%22Med_Mdcr_Alowd_Amt%22%2C%22Med_Mdcr_Pymt_Amt%22%2C%22Med_Mdcr_Stdzd_Amt%22%5D%7D

2015

https://data.cms.gov/provider-summary-by-type-of-service/medicare-physician-other-practitioners/medicare-physician-other-practitioners-by-provider/data/2015?query=%7B%22filters%22%3A%7B%22list%22%3A%5B%7B%22conjunction%22%3A%7B%22value%22%3A%22OR%22%7D%2C%22conditions%22%3A%5B%7B%22column%22%3A%7B%22value%22%3A%22Rndrng_Prvdr_Type%22%7D%2C%22comparator%22%3A%7B%22value%22%3A%22%3D%22%7D%2C%22filterValue%22%3A%5B%22anesthesiology%22%5D%7D%2C%7B%22column%22%3A%7B%22value%22%3A%22Rndrng_Prvdr_Type%22%7D%2C%22comparator%22%3A%7B%22value%22%3A%22%3D%22%7D%2C%22filterValue%22%3A%5B%22CRNA%22%5D%7D%2C%7B%22column%22%3A%7B%22value%22%3A%22Rndrng_Prvdr_Type%22%7D%2C%22comparator%22%3A%7B%22value%22%3A%22%3D%22%7D%2C%22filterValue%22%3A%5B%22anesthesiologist%20assistants%22%5D%7D%5D%7D%5D%2C%22rootConjunction%22%3A%7B%22value%22%3A%22AND%22%7D%7D%2C%22keywords%22%3A%22%22%2C%22offset%22%3A0%2C%22limit%22%3A10%2C%22sort%22%3A%7B%22sortBy%22%3Anull%2C%22sortOrder%22%3Anull%7D%2C%22columns%22%3A%5B%22Rndrng_Prvdr_Type%22%2C%22Rndrng_Prvdr_State_Abrvtn%22%2C%22Tot_HCPCS_Cds%22%2C%22Tot_Benes%22%2C%22Tot_Srvcs%22%2C%22Tot_Sbmtd_Chrg%22%2C%22Tot_Mdcr_Alowd_Amt%22%2C%22Tot_Mdcr_Pymt_Amt%22%2C%22Tot_Mdcr_Stdzd_Amt%22%2C%22Med_Tot_HCPCS_Cds%22%2C%22Med_Tot_Benes%22%2C%22Med_Tot_Srvcs%22%2C%22Med_Sbmtd_Chrg%22%2C%22Med_Mdcr_Alowd_Amt%22%2C%22Med_Mdcr_Pymt_Amt%22%2C%22Med_Mdcr_Stdzd_Amt%22%5D%7D

2016

https://data.cms.gov/provider-summary-by-type-of-service/medicare-physician-other-practitioners/medicare-physician-other-practitioners-by-provider/data/2016?query=%7B%22filters%22%3A%7B%22list%22%3A%5B%7B%22conjunction%22%3A%7B%22value%22%3A%22OR%22%7D%2C%22conditions%22%3A%5B%7B%22column%22%3A%7B%22value%22%3A%22Rndrng_Prvdr_Type%22%7D%2C%22comparator%22%3A%7B%22value%22%3A%22%3D%22%7D%2C%22filterValue%22%3A%5B%22anesthesiology%22%5D%7D%2C%7B%22column%22%3A%7B%22value%22%3A%22Rndrng_Prvdr_Type%22%7D%2C%22comparator%22%3A%7B%22value%22%3A%22%3D%22%7D%2C%22filterValue%22%3A%5B%22certified%20registered%20nurse%20anesthetist%20%28CRNA%29%22%5D%7D%2C%7B%22column%22%3A%7B%22value%22%3A%22Rndrng_Prvdr_Type%22%7D%2C%22comparator%22%3A%7B%22value%22%3A%22%3D%22%7D%2C%22filterValue%22%3A%5B%22anesthesiology%20assistant%22%5D%7D%5D%7D%5D%2C%22rootConjunction%22%3A%7B%22value%22%3A%22AND%22%7D%7D%2C%22keywords%22%3A%22%22%2C%22offset%22%3A0%2C%22limit%22%3A10%2C%22sort%22%3A%7B%22sortBy%22%3Anull%2C%22sortOrder%22%3Anull%7D%2C%22columns%22%3A%5B%22Rndrng_Prvdr_Type%22%2C%22Rndrng_Prvdr_State_Abrvtn%22%2C%22Tot_HCPCS_Cds%22%2C%22Tot_Benes%22%2C%22Tot_Srvcs%22%2C%22Tot_Sbmtd_Chrg%22%2C%22Tot_Mdcr_Alowd_Amt%22%2C%22Tot_Mdcr_Pymt_Amt%22%2C%22Tot_Mdcr_Stdzd_Amt%22%2C%22Med_Tot_HCPCS_Cds%22%2C%22Med_Tot_Benes%22%2C%22Med_Tot_Srvcs%22%2C%22Med_Sbmtd_Chrg%22%2C%22Med_Mdcr_Alowd_Amt%22%2C%22Med_Mdcr_Pymt_Amt%22%2C%22Med_Mdcr_Stdzd_Amt%22%5D%7D

2017

https://data.cms.gov/provider-summary-by-type-of-service/medicare-physician-other-practitioners/medicare-physician-other-practitioners-by-provider/data/2017?query=%7B%22filters%22%3A%7B%22list%22%3A%5B%7B%22conjunction%22%3A%7B%22value%22%3A%22OR%22%7D%2C%22conditions%22%3A%5B%7B%22column%22%3A%7B%22value%22%3A%22Rndrng_Prvdr_Type%22%7D%2C%22comparator%22%3A%7B%22value%22%3A%22%3D%22%7D%2C%22filterValue%22%3A%5B%22anesthesiology%22%5D%7D%2C%7B%22column%22%3A%7B%22value%22%3A%22Rndrng_Prvdr_Type%22%7D%2C%22comparator%22%3A%7B%22value%22%3A%22%3D%22%7D%2C%22filterValue%22%3A%5B%22certified%20registered%20nurse%20anesthetist%20%28CRNA%29%22%5D%7D%2C%7B%22column%22%3A%7B%22value%22%3A%22Rndrng_Prvdr_Type%22%7D%2C%22comparator%22%3A%7B%22value%22%3A%22%3D%22%7D%2C%22filterValue%22%3A%5B%22anesthesiology%20assistant%22%5D%7D%5D%7D%5D%2C%22rootConjunction%22%3A%7B%22value%22%3A%22AND%22%7D%7D%2C%22keywords%22%3A%22%22%2C%22offset%22%3A0%2C%22limit%22%3A10%2C%22sort%22%3A%7B%22sortBy%22%3Anull%2C%22sortOrder%22%3Anull%7D%2C%22columns%22%3A%5B%22Rndrng_Prvdr_Type%22%2C%22Rndrng_Prvdr_State_Abrvtn%22%2C%22Tot_HCPCS_Cds%22%2C%22Tot_Benes%22%2C%22Tot_Srvcs%22%2C%22Tot_Sbmtd_Chrg%22%2C%22Tot_Mdcr_Alowd_Amt%22%2C%22Tot_Mdcr_Pymt_Amt%22%2C%22Tot_Mdcr_Stdzd_Amt%22%2C%22Med_Tot_HCPCS_Cds%22%2C%22Med_Tot_Benes%22%2C%22Med_Tot_Srvcs%22%2C%22Med_Sbmtd_Chrg%22%2C%22Med_Mdcr_Alowd_Amt%22%2C%22Med_Mdcr_Pymt_Amt%22%2C%22Med_Mdcr_Stdzd_Amt%22%5D%7D

2018

https://data.cms.gov/provider-summary-by-type-of-service/medicare-physician-other-practitioners/medicare-physician-other-practitioners-by-provider/data/2018?query=%7B%22filters%22%3A%7B%22list%22%3A%5B%7B%22conjunction%22%3A%7B%22value%22%3A%22OR%22%7D%2C%22conditions%22%3A%5B%7B%22column%22%3A%7B%22value%22%3A%22Rndrng_Prvdr_Type%22%7D%2C%22comparator%22%3A%7B%22value%22%3A%22%3D%22%7D%2C%22filterValue%22%3A%5B%22anesthesiology%22%5D%7D%2C%7B%22column%22%3A%7B%22value%22%3A%22Rndrng_Prvdr_Type%22%7D%2C%22comparator%22%3A%7B%22value%22%3A%22%3D%22%7D%2C%22filterValue%22%3A%5B%22certified%20registered%20nurse%20anesthetist%20%28CRNA%29%22%5D%7D%2C%7B%22column%22%3A%7B%22value%22%3A%22Rndrng_Prvdr_Type%22%7D%2C%22comparator%22%3A%7B%22value%22%3A%22%3D%22%7D%2C%22filterValue%22%3A%5B%22anesthesiology%20assistant%22%5D%7D%5D%7D%5D%2C%22rootConjunction%22%3A%7B%22value%22%3A%22AND%22%7D%7D%2C%22keywords%22%3A%22%22%2C%22offset%22%3A0%2C%22limit%22%3A10%2C%22sort%22%3A%7B%22sortBy%22%3Anull%2C%22sortOrder%22%3Anull%7D%2C%22columns%22%3A%5B%22Rndrng_Prvdr_Type%22%2C%22Rndrng_Prvdr_State_Abrvtn%22%2C%22Tot_HCPCS_Cds%22%2C%22Tot_Benes%22%2C%22Tot_Srvcs%22%2C%22Tot_Sbmtd_Chrg%22%2C%22Tot_Mdcr_Alowd_Amt%22%2C%22Tot_Mdcr_Pymt_Amt%22%2C%22Tot_Mdcr_Stdzd_Amt%22%2C%22Med_Tot_HCPCS_Cds%22%2C%22Med_Tot_Benes%22%2C%22Med_Tot_Srvcs%22%2C%22Med_Sbmtd_Chrg%22%2C%22Med_Mdcr_Alowd_Amt%22%2C%22Med_Mdcr_Pymt_Amt%22%2C%22Med_Mdcr_Stdzd_Amt%22%5D%7D

2019

https://data.cms.gov/provider-summary-by-type-of-service/medicare-physician-other-
practitioners/medicare-physician-other-practitioners-by-
provider/data/2019?query=%7B%22filters%22%3A%7B%22list%22%3A%5B%7B%22conjunction%22%3A
%7B%22value%22%3A%22OR%22%7D%2C%22conditions%22%3A%5B%7B%22column%22%3A%7B%22
value%22%3A%22Rndrng_Prvdr_Type%22%7D%2C%22comparator%22%3A%7B%22value%22%3A%22
%3D%22%7D%2C%22filterValue%22%3A%5B%22anesthesiology%22%5D%7D%2C%7B%22column%22%
3A%7B%22value%22%3A%22Rndrng_Prvdr_Type%22%7D%2C%22comparator%22%3A%7B%22value%2
2%3A%22%3D%22%7D%2C%22filterValue%22%3A%5B%22certified%20registered%20nurse%20anesthe
tist%20%28CRNA%29%22%5D%7D%2C%7B%22column%22%3A%7B%22value%22%3A%22Rndrng_Prvd
r_Type%22%7D%2C%22comparator%22%3A%7B%22value%22%3A%22%3D%22%7D%2C%22filterValue
%22%3A%5B%22anesthesiology%20assistant%22%5D%7D%5D%7D%5D%2C%22rootConjunction%22%3
A%7B%22value%22%3A%22AND%22%7D%7D%2C%22keywords%22%3A%22%22%2C%22offset%22%3A
0%2C%22limit%22%3A10%2C%22sort%22%3A%7B%22sortBy%22%3Anull%2C%22sortOrder%22%3Anul
l%7D%2C%22columns%22%3A%5B%22Rndrng_Prvdr_Type%22%2C%22Rndrng_Prvdr_State_Abrvtn%22
%2C%22Tot_HCPCS_Cds%22%2C%22Tot_Benes%22%2C%22Tot_Srvcs%22%2C%22Tot_Sbmtd_Chrg%22
%2C%22Tot_Mdcr_Alowd_Amt%22%2C%22Tot_Mdcr_Pymt_Amt%22%2C%22Tot_Mdcr_Stdzd_Amt%2
2%2C%22Med_Tot_HCPCS_Cds%22%2C%22Med_Tot_Benes%22%2C%22Med_Tot_Srvcs%22%2C%22M
ed_Sbmtd_Chrg%22%2C%22Med_Mdcr_Alowd_Amt%22%2C%22Med_Mdcr_Pymt_Amt%22%2C%22M
ed_Mdcr_Stdzd_Amt%22%5D%7D

2020

https://data.cms.gov/provider-summary-by-type-of-service/medicare-physician-other-practitioners/medicare-physician-other-practitioners-by-provider/data/2020?query=%7B%22filters%22%3A%7B%22list%22%3A%5B%7B%22conjunction%22%3A%7B%22value%22%3A%22OR%22%7D%2C%22conditions%22%3A%5B%7B%22column%22%3A%7B%22value%22%3A%22Rndrng_Prvdr_Type%22%7D%2C%22comparator%22%3A%7B%22value%22%3A%22%3D%22%7D%2C%22filterValue%22%3A%5B%22anesthesiology%22%5D%7D%2C%7B%22column%22%3A%7B%22value%22%3A%22Rndrng_Prvdr_Type%22%7D%2C%22comparator%22%3A%7B%22value%22%3A%22%3D%22%7D%2C%22filterValue%22%3A%5B%22certified%20registered%20nurse%20anesthetist%20%28CRNA%29%22%5D%7D%2C%7B%22column%22%3A%7B%22value%22%3A%22Rndrng_Prvdr_Type%22%7D%2C%22comparator%22%3A%7B%22value%22%3A%22%3D%22%7D%2C%22filterValue%22%3A%5B%22anesthesiology%20assistant%22%5D%7D%5D%7D%5D%2C%22rootConjunction%22%3A%7B%22value%22%3A%22AND%22%7D%7D%2C%22keywords%22%3A%22%22%2C%22offset%22%3A0%2C%22limit%22%3A10%2C%22sort%22%3A%7B%22sortBy%22%3Anull%2C%22sortOrder%22%3Anull%7D%2C%22columns%22%3A%5B%22Rndrng_Prvdr_Type%22%2C%22Rndrng_Prvdr_State_Abrvtn%22%2C%22Tot_HCPCS_Cds%22%2C%22Tot_Benes%22%2C%22Tot_Srvcs%22%2C%22Tot_Sbmtd_Chrg%22%2C%22Tot_Mdcr_Alowd_Amt%22%2C%22Tot_Mdcr_Pymt_Amt%22%2C%22Tot_Mdcr_Stdzd_Amt%22%2C%22Med_Tot_HCPCS_Cds%22%2C%22Med_Tot_Benes%22%2C%22Med_Tot_Srvcs%22%2C%22Med_Sbmtd_Chrg%22%2C%22Med_Mdcr_Alowd_Amt%22%2C%22Med_Mdcr_Pymt_Amt%22%2C%22Med_Mdcr_Stdzd_Amt%22%5D%7D

2021

https://data.cms.gov/provider-summary-by-type-of-service/medicare-physician-other-practitioners/medicare-physician-other-practitioners-by-provider/data/2021?query=%7B%22filters%22%3A%7B%22list%22%3A%5B%7B%22conjunction%22%3A%7B%22value%22%3A%22OR%22%7D%2C%22conditions%22%3A%5B%7B%22column%22%3A%7B%22value%22%3A%22Rndrng_Prvdr_Type%22%7D%2C%22comparator%22%3A%7B%22value%22%3A%22%3D%22%7D%2C%22filterValue%22%3A%5B%22anesthesiology%22%5D%7D%2C%7B%22column%22%3A%7B%22value%22%3A%22Rndrng_Prvdr_Type%22%7D%2C%22comparator%22%3A%7B%22value%22%3A%22%3D%22%7D%2C%22filterValue%22%3A%5B%22certified%20registered%20nurse%20anesthetist%20%28CRNA%29%22%5D%7D%2C%7B%22column%22%3A%7B%22value%22%3A%22Rndrng_Prvdr_Type%22%7D%2C%22comparator%22%3A%7B%22value%22%3A%22%3D%22%7D%2C%22filterValue%22%3A%5B%22anesthesiology%20assistant%22%5D%7D%5D%7D%5D%2C%22rootConjunction%22%3A%7B%22value%22%3A%22AND%22%7D%7D%2C%22keywords%22%3A%22%22%2C%22offset%22%3A0%2C%22limit%22%3A10%2C%22sort%22%3A%7B%22sortBy%22%3Anull%2C%22sortOrder%22%3Anull%7D%2C%22columns%22%3A%5B%22Rndrng_Prvdr_Type%22%2C%22Rndrng_Prvdr_State_Abrvtn%22%2C%22Tot_HCPCS_Cds%22%2C%22Tot_Benes%22%2C%22Tot_Srvcs%22%2C%22Tot_Sbmtd_Chrg%22%2C%22Tot_Mdcr_Alowd_Amt%22%2C%22Tot_Mdcr_Pymt_Amt%22%2C%22Tot_Mdcr_Stdzd_Amt%22%2C%22Med_Tot_HCPCS_Cds%22%2C%22Med_Tot_Benes%22%2C%22Med_Tot_Srvcs%22%2C%22Med_Sbmtd_Chrg%22%2C%22Med_Mdcr_Alowd_Amt%22%2C%22Med_Mdcr_Pymt_Amt%22%2C%22Med_Mdcr_Stdzd_Amt%22%5D%7D
